# Which policies most effectively reduce SARS-CoV-2 transmission in schools?

**DOI:** 10.1101/2020.11.24.20237305

**Authors:** Anna Bershteyn, Hae-Young Kim, Jessica McGillen, R. Scott Braithwaite

## Abstract

**Introduction:** New York City (NYC) has the largest public school system in the United States (US). During the SARS-CoV-2 pandemic, NYC was the first major US city to open schools for in-person learning in the 2020-2021 academic year. Several policies were implemented to reduce the risk of in-school transmission, including infection control measures (facemasks, physical distancing, enhanced indoor ventilation, cohorting of small groups, and hand hygiene), option of all-remote instruction, alternative options for how class schedules would rotate in-person and remote instruction, daily symptom screening, and testing 10-20% of students and staff weekly or monthly depending on local case rates. We sought to determine which of these policies had the greatest impact on reducing the risk of in-school transmission.

**Methods:** We evaluated the impact of each policy by referring to global benchmarks for the secondary attack rate (SAR) of SARS-CoV-2 in school settings and by simulating the potential for transmission in NYC’s rotating cohort schedules, in which teachers could act as “bridges” across rotating cohorts. We estimated the impact of (1) infection control measures, (2) providing an option of all-remote instruction, (3) choice of class scheduling for in-person learners, (4) daily symptom screening, (5) testing to curtail transmission, and (6) testing to identify school outbreaks. Each policy was assessed independently of other policies, with the exception of symptom screening and random testing, which were assessed both independently and jointly.

**Results:** Among the policies analyzed, the greatest transmission reduction was associated with the infection control measures, followed by small class cohorts with an option for all-remote instruction, symptom screening, and finally randomly testing 10-20% of school attendees. Assuming adult staff are the primary source of within-school SARS-CoV-2 transmission, weekly testing of staff could be at least as effective as symptom screening, and potentially more so if testing days occur in the beginning of the workweek with results available by the following day. A combination of daily symptom screening and testing on the first workday of each week could reduce transmission by 70%.

**Conclusions:** Adherence to infection control is the highest priority for safe school re-opening. Further transmission reduction can be achieved through small rotating class cohorts with an option for remote learning, widespread testing at the beginning of the work week, and daily symptom screening and self-isolation. Randomly testing 10-20% of attendees weekly or monthly does not meaningfully curtail transmission and may not detect outbreaks before they have spread beyond a handful of individuals. School systems considering re-opening during the SARS-CoV-2 pandemic or similarly virulent respiratory disease outbreaks should consider these relative impacts when setting policy priorities.

## Introduction

New York City (NYC), with the largest school district in the United States, was the first major US city to offer in-person instruction during the 2020-2021 school year despite persistent community transmission after the first SARS-CoV-2 pandemic wave. Several policies were implemented to adapt education to the pandemic context.

First, schools implemented infection control measures related to individual behavior as well as the use of school infrastructure. Individuals were required to wear face coverings, maintain at least 6 feet of physical distance from others, and wash hands or use hand sanitizer.^1^ Buildings were inspected to ensure that all occupied rooms had at least one operable window, mechanical exhaust fan, supply fan, or unit ventilator delivering airflow to the space.^2^ The number of contacts per individual were minimized by subdividing classes into cohorts of 9 to 12 students attending on school alternate days, canceling large gatherings such as assemblies, and consuming meals in classrooms rather than cafeterias.

Second, while cohorting is fundamentally a part of infection control, the NYC school system implemented a unique policy by which school principals could select one of six schedules for cohorts to rotate between in-person and remote instruction. Two groups of up to 13 students could attend on alternating Monday and on two designated days, either staggered (Tuesday and Thursday, or Wednesday and Friday) or in two-day blocks (Tuesday and Wednesday, or Thursday and Friday). Three groups of 9 students could attend on one designated day, and then on one day every two out of three weeks, or else one two-day block once every three weeks. Finally, for older students, three groups of 9 students could attend in a 6-day rotation either once every three days, or two consecutive days in every six days. We evaluated whether the choice of cohort schedule impacted the risk of transmission of SARS-CoV-2, considering that teachers could act as transmission “bridges” across cohorts.

Third, potential SARS-CoV-2 infections were monitored through symptom screening and in-school testing. Prior to arriving at school, attendees were required to complete an online or paper questionnaire on whether they had experienced fever >100F, new cough, new loss of test or smell, or shortness of breath in the preceding 10 days.^3^ Symptomatic individuals must self-isolate for 10 days unless they had received a negative SARS-CoV-2 viral test and been symptom-free for at least 24 hours. Testing was provided in school buildings, either monthly or weekly depending on whether neighborhood rates of SARS-CoV-2 were above or below 2.5 per 100,000 residents, to a randomly selected 10% to 20% of school students and staff. While this testing program primarily served as a form of surveillance assess the prevalence of SARS-CoV-2 infection in schools, here we assess its direct effect on transmission reduction and calculate the potential effect of even more frequent in-school testing.

Finally, school closure policies were established according to the number of positive viral detected at a school. Detection of a single case would lead to a two-week quarantine of the class cohort, instructor(s), and any other close contacts of the index case, while detection of two or more cases in the same school would trigger a two-week closure of the entire school, unless the two cases were linked by known contact (for example, siblings sharing a household or classmates in the same cohort). While these policies are reactive, rather than preventative, of initial in-school transmission, they may prevent the spread of SARS-CoV-2 beyond a handful of individuals if an outbreak is detected early. We evaluated the size of an outbreak that could be detected by 10% or 20% in-school testing.

We evaluated the impact of each policy measure in NYC schools on SARS-CoV-2 transmission by synthesizing NYC and global school re-opening data and simulating in-school outbreaks under different cohort rotation and testing scenarios. The goal of the analysis was to identify which components of the NYC model contributed most to student and staff safety during the SARS-CoV-2 pandemic.

## Methods

### Infection control measures

In order to identify in-school transmission rates measured independently from the rate of case importation into schools, we identified studies of the secondary attack rate (SAR) of SARS-CoV-2 in schools that had a confirmed positive index case and were conducting in-person instruction prior to NYC school re-opening.

We identified one report estimating SAR in the absence of infection control. In Israeli schools, 13.1% of attendees tested positive among 1,352 attendees tested.^4^ Schools remained open for only ten days. This SAR is within the range of observed within-household SAR for SARS-CoV-2, suggesting that sharing a classroom may harbor a similar level of risk to sharing a household in the absence of infection control. Household transmission studies have reported an SAR of 18.1% (range across studies: 3.9%—54 .9%),^5^ which in this analysis we assumed to be a worst-case SAR in the absence of infection control measures.

Partial implementation of measures in New South Wales – for example, masks worn by adults but not young children – occurred while 27 index cases comprised of 15 adults and 12 children attended schools with 1,448 other individuals, of whom 633 were tested and 18 tested positive, for an estimated SAR of 1.4% – 2.8%.

Fuller implementation of facemasks, physical distancing, ventilation, cohorting, and hand hygiene were implemented in South Korea and Germany. In Saxony, Germany, an SAR study among school attendees measured antibodies against SARS-CoV-2, a longer-lasting biomarker that does not necessarily indicate current infection, and therefore may provide an upper bound to SAR in the context of infection control measures. In the study, 0.6% of attendees tested positive for SARS-CoV-2 antibodies with no clusters exceeding 4 individuals.^6^ In South Korea, 4,768 individuals who attended schools with 15 index cases received SARS-CoV-2 viral testing, which indicates current infection. Only one individual tested positive, for an SAR of 0.021%.^7^

We compared these SARs to each other in order to assess the potential impact of fully implemented infection control measures. We also compared these SARs to the median enrollment in NYC public schools prior to the SARS-CoV-2 pandemic – 475 students – in order to assess whether infection control would be sufficient to prevent outbreaks in schools.

### Option of remote instruction

While establishing small cohorts may require students have partially remote learning due to limited building capacity and teachers, this does not necessarily require a separate option for students to choose fully-remote learning. For this reason, our analysis separately examines the impact of the fully-remote learning option. We based our analysis on media reports providing the fraction of NYC students who opted for fully-remote versus hybrid (remote and in-person) learning as of October 13, 2020.^8^

### Choice of class schedule

We developed a simulation model to compare the number of secondary infections that could arise from a teacher acting as a bridge across rotating cohorts in a hypothetical scenario where the classroom environment is conducive to transmission, e.g., due to non-adherence to infection control measures.

Cohort rotation schedules available from NYC DOE^9^ were simulated with the initial condition of a single infected teacher working five days per week with two or three cohorts, depending on the schedule. We assumed the teacher had equal probability of becoming infected any day of the week, assuming worst-case-scenario school-day SAR equal to the household SAR of 18.1% (3.9%—54 .9%)representing the absence of concurrent infection control measures.^5^ We distributed the transmission from the index case over two weeks, with transmission varying according to a gamma distribution 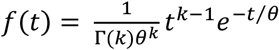 with a shape parameter *k* = 2.25 and a scale parameter *θ* = 2.80. The distribution was chosen because it mirrors the growth-and-fall of viral shedding viral shedding, peaking on the 4^th^ and 5^th^ day of infection,^10,11^ and the parameters were selected to match the distribution of observed serial intervals for SARS-CoV-2 transmission.^12^

### Daily symptom screening

Daily symptom screening consists of temperature checks and qualitative self-assessment of symptoms prior to entry into school buildings. Symptomatic individuals are asked to self-isolate for 10 days and until symptom-free for at least 24 hours. We assumed that 69% (61%—74%) of index cases would develop symptoms on the fifth day of infection based on a meta-analysis of SARS-CoV-2 symptom development.^13^ and would self-isolate on subsequent days. For each NYC DOE cohort rotation schedule, we simulated estimated the impact of testing and symptom screening for curtailing SARS-COV-2 transmission. s

### Testing to curtail transmission

For each NYC DOE cohort rotation schedule, we simulated different levels of viral testing – monthly or weekly with 10%, 20%, or 100% probability – assuming that the index case would test positive on days when they were infectious and that test results would be returned by the following day, so that the index case may transmit on the day of testing but not on subsequent days.

### Testing to identify school outbreaks

We represented outbreak detection as reactive strategy that would not reduce the probability of transmission from an index case, but may limit the size of an outbreak by interrupting transmission before outbreaks spread beyond a handful of people. We used random sampling without replacement to estimate the size of an outbreak that could be detected by random testing during the outbreak with ≥50% probability (more likely than not to detect the outbreak) and ≥90% probability (would reliably detect the outbreak) in the median school population among NYC public schools, after adjusting for those opting for fully-remote learning.

## Results

### Infection control measures

The SAR in Israeli school re-opening in the absence of infection control was 625-fold higher than the SAR of South Korean schools in which infection control was implemented. The measurement of SAR in Saxony where infection control was implemented, which may be an overestimate due to the use antibody testing rather than viral testing, was 21.8-fold higher than the SAR in Israel. No other intervention tested in this study had as large an effect size on SARS-CoV-2 transmission.

### Option of remote instruction

NYC provided parents with a choice between all-remote instruction and a hybrid of in-person and remote instruction. Half of students opted for all-remote instruction as of October 2020. The option reduced by up to half the number potential index cases who unknowingly attend school while infected, and also reduced by half the number of susceptible individuals attending schools, for an overall transmission reduction of up to 75%. We note that the effect may be more modest if students who opt-out are those who are also likely to acquire SARS-CoV-2 (for example, people who are medically vulnerable or have vulnerable household members and therefore may take greater precautions) but were unable to quantify the differential infection risk among those opting for or against in-person instruction.

### Choice of class scheduling

NYC school principals could select among six schedules in which students could attend either half or one-third of school days in-person.^9^ In simulating the potential secondary spread from a teacher acting as a “bridge” across rotating cohorts (Figure 1), we found that smaller cohorts and commensurately reduced instruction time – cohorts of 9 attending one-third of days (orange bars) rather than cohorts of 13 attending one-half of days (blue bars) – reduced transmission risk However, for a given cohort size, the choice of rotation schedule among those on the approved list had minimal impact on transmission.

**Figure 1.**
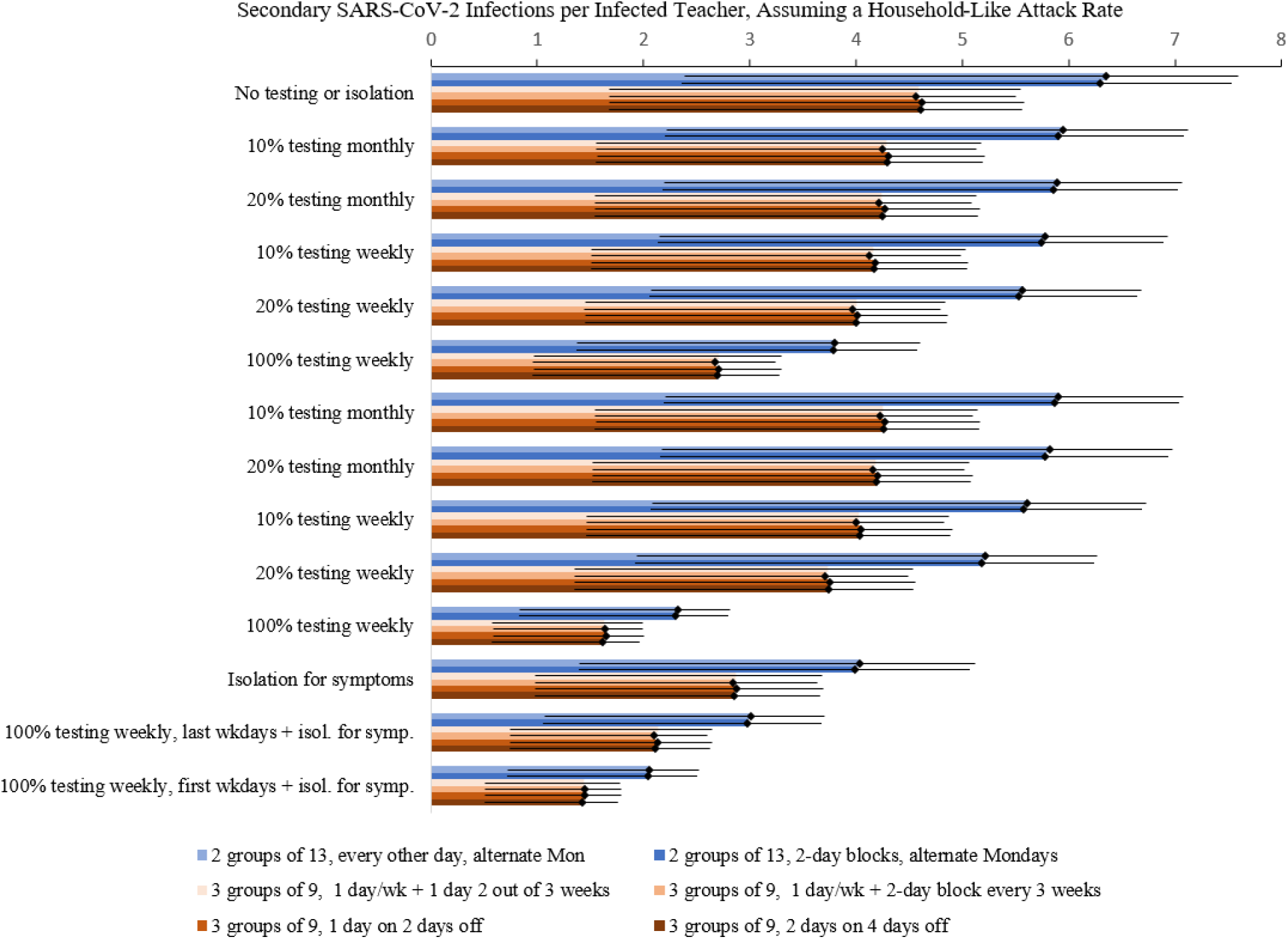
Simulated classroom spread of SARS-CoV-2 from an infected teacher, assuming SAR based on household transmission studies. Colored bars represent each of 6 classroom rotation models available for NYC Public Schools. Policies include daily symptom screening, and monthly or weekly testing of 10%, 20%, or 100% of attendees, with testing occurring either on the most optimal day (the first weekday of a 5-day work week, which is Monday for US public schools) or the least optimal day (the last weekday of a 5-day week, which is Friday for US public schools). Daily symptom screening alone is estimated to reduce transmission by 34.8-41.8%, more than 10% or 20% testing and on par with weekly 100% testing on the least optimal day (the last weekday). Weekly 100% testing on the most optimal day (the first weekday) without symptom screening eliminates three-fifths of transmission. The combination of weekly testing on the first weekday and symptom screening would have the greatest impact, reducing transmission by 70%.

### Daily symptom screening

With a policy requiring index cases to self-isolate if they develop symptoms, in-school transmission only occurs during pre-symptomatic infection (days 1 through 4) and asymptomatic infection (26 to 39% of index cases^13^). In the absence of additional testing for asymptomatic individuals, this policy reduces transmission by 34.8-41.8% relative to no isolation (Figure 1).

### Testing to curtail transmission

NYC’s school SARS-CoV-2 testing program consisting of monthly or weekly testing of 10% or 20% of in-school students and staff selected at random. The impact of this testing on transmission from an infected index case was modest (Figure 1) due to the low probability of identifying the index case prior to onward transmission. We therefore also simulated a policy of universal weekly testing, which would identify 100% of index cases and recur on a designated day of the week.

The impact of weekly testing depended on the day of the week in which testing was deployed, due to the lack of in-school transmission over the two-day weekend. Assuming test results could be returned by the next day, we found that the first weekday (Mondays in US schools) was the most optimal day for testing, while the last weekday (Fridays in US schools) was the least optimal. Testing on Monday averted 27.1-34.0% more infections than testing on Friday and could reduce transmission by 61.8-64.2% without symptom-based isolation.

The most effective testing and isolation strategy used a combination of 100% testing on the first weekday together with symptom screening and isolation of all those who are symptomatic, for an overall transmission reduction of 68.6-71.1% relative to no testing or symptom-based screening.

### Testing to identify school outbreaks

Random testing of 10% to 20% of in-school attendees may also identify secondary spread of SARS-CoV-2 infection and precipitate outbreak investigation and school closure. In an average NYC school size across of 339 weekly in-person attendees (after adjusting for those opting for fully-remote learning), detection of a at least one positive case through 20% random testing would require the outbreak to grow to at least 4 and 11 positive individuals in order to be detected with >50% and >90% probability, respectively. With 10% random testing, the outbreak would need to grow to 7 and 22 cases in order for the first case to be detected with >50% and >90% probability, respectively.

While a single positive case would trigger quarantine of the case’s close contacts, including classmates in the cohort and instructor(s), detection of two or more cases would trigger a school closure and investigation of a possible a school-wide outbreak. Outbreak detection (of two or more cases) through 20% random testing would require the outbreak to grow to at least 8 and 18 positive individuals in order to be detected with >50% and >90% probability, respectively. With 10% random testing, the outbreak would need to grow to 17 and 36 cases in order for the outbreak to be detected with >50% and >90% probability, respectively.

## Discussion

Several policies were implemented as part of the re-opening of NYC Public Schools during the SARS-CoV-2 pandemic. We assessed which policies were likely to have been most effective at reducing SARS-CoV-2 transmission in the school environment. We found that infection control measures (facemasks, physical distancing, hand hygiene, and room ventilation) may contribute more than any other policy to the prevention of SARS-CoV-2 transmission in schools.

After infection control, the next most effective policy is to provide an option for all-remote instruction, particularly in populations such as New York City where approximately half of students opt to learn remotely on a full-time basis. This policy provides a dual benefit for transmission reduction: first, it reduces the number of potential index cases in the school environment at any given time, and second, it reduces the number of potential contacts an index case may expose while attending school. A limitation of this policy is that policy-makers cannot control and may struggle to anticipate the fraction of students who will opt for fully-remote instruction.

The specific rotation schedule had little impact on the risk of transmission, even if a teacher were to become infected and act as a “bridge” across cohorts. There was greater transmission risk in rotation schedules that involved larger cohorts with more days of in-person attendance per student, but these effects were modest compared to the large impact of infection control and providing an option of fully-remote learning. These findings suggest that schools should select the rotation schedule most conducive to their education mission, while focusing on infection control as a primary means of transmission prevention.

Testing and symptom screening may have a moderate contribution to reducing transmission even under optimistic assumptions. We found that daily symptom screening could reduce transmission by one-third to two-fifths, while NYC’s strategy of testing 10-20% of school attendees had little direct effect on transmission reduction, nor could these testing rates reliably detect outbreaks before they spread beyond a handful of individuals. At these rates of in-school testing, detection of smaller outbreaks depends upon the initiative of students and staff to seek outside testing at health facilities and test centers. In NYC, testing was free and unrestricted throughout the fall of 2020; other school districts should consider the availability of outside testing when deciding whether outside testing can contribute to outbreak detection among school attendees.

Increasing rates of in-school testing could amplify its impact until it is commensurate with the next-best policy option (the choice of all-remote learning). If schools could increase testing rates to cover 100% of on weekly the first weekday, with results available by the following day, testing alone could reduce transmission by approximately three-fifths. The combination of weekly testing on the first weekday and symptom screening would have the greatest impact, reducing transmission by approximately 70% – a transmission reduction comparable to the 75% reduction from the fully-remote learning option.

### Limitations

Our study has several limitations. We did not conduct a formal systematic review of the effect of infection control and other policies within schools due to the paucity of published and controlled studies in the first year of the SARS-CoV-2 pandemic. We relied on global benchmarks of SAR from school systems that conducted widespread testing after identifying index cases in school systems. These global experiences were not standardized in terms of the ages and grades of the students, the details of how each infection control measure was implemented (e.g., mask type, type of ventilation, minimal physical distance, use of physical barriers), the type of specimen and test used to assess secondary spread, or the context surrounding the outbreak such as the climate and level of community transmission at the time of the outbreak. For this reason, we could not quantitatively estimate the impact of infection control, beyond the observation that SAR was orders of magnitude lower in studies conducted where infection control had been implemented. However, the large magnitude of difference clearly exceeded the more precisely quantifiable effects of policies such as testing and cohort rotation, and is consistent with observational evidence from non-educational settings that maintaining six feet of distance from an index case can reduce transmission by four-fold compared to close physical proximity, while facemask use can further reduce transmission by approximately seven-fold.^14^

Our study did not investigate the potential for differential testing strategies by student age or grade. Preliminary evidence suggests a greater SARS-CoV-2 transmission and acquisition potential among adolescents and teenagers cosmpared to younger children.^15^ Therefore, our classroom rotation analysis, which focuses on teachers as a bridge between class cohorts, may be most applicable to younger grades for whom teachers are the most likely to acquire and transmit SARS-CoV-2.

Finally, our study did not consider school-related contacts outside of classroom cohorts, such as contacts related to transportation, staff meetings or trainings, or after-school care. Further study is needed to determine how in-person education influences the number of contacts among students and staff beyond the classroom in the context of a pandemic.

## Conclusions

NYC school re-opening has the potential to inform policies in other school systems, as well as policies to be adopted in future respiratory disease epidemics. As evidence accrues about the effectiveness of school re-opening policies, it is critical that the right lessons are learned and that the most effective policies are prioritized to ensure that education can proceed safely.

Adherence to in-school infection control measures including facemasks, physical distancing, enhanced room ventilation, cohorting, and hand hygiene reduced in-school transmission rates by multiple orders of magnitude and are the most effective policy. Providing an option of remote learning reduced transmission substantially, by up to three-quarters relative to mandatory in-person education. Weekly testing at the beginning of the workweek combined with symptom screening and isolation could reduce transmission by 70%, while symptom screening alone reduces transmission by 33-42%. Lower levels of testing, such as the weekly or monthly 10-20% random testing conducted in NYC, has minimal direct impact on transmission. We conclude that adherence to infection control is the highest priority for safe school re-opening, and that further transmission reduction can be achieved by providing a remote learning option, widespread testing at the beginning of the work week, and daily symptom screening and self-isolation.School systems considering re-opening during the SARS-CoV-2 pandemic or similar respiratory disease outbreaks should consider these relative impacts when setting policy priorities.

## Data Availability

Only publicly available data were used in this manuscript. Links to data sources have been referenced in the manuscript.

